# Dietary Iron Intake and Mental and Behavioral Disorders Due to Use of Tobacco: A Cross-Sectional and Longitudinal Study Based on the UK Biobank

**DOI:** 10.1101/2024.10.30.24316458

**Authors:** Xueting Qi, Ronghui Zhang, Hailong Zhu, Jia Luo, Qiuge Zhang, Weijing Wang, Tong Wang, Dongfeng Zhang

**Author notes:** **Corresponding author:** Dongfeng Zhang, Department of Epidemiology and Health Statistics, the School of Public Health of Qingdao University, 308 Ningxia Road, Qingdao, Shandong 266071, People’s Republic of China., Tong Wang, Department of Epidemiology and Health Statistics, the School of Public Health of Qingdao University, 308 Ningxia Road, Qingdao, Shandong 266071, People’s Republic of China.

## Abstract

**Introduction:** Over 1 billion smokers worldwide, one-third of whom have mental and behavioral disorders, exist. However, factors influencing mental and behavioral disorders due to use of tobacco remain unexplored. We aim to investigate the relationship between dietary iron intake and mental and behavioral disorders due to use of tobacco.

**Methods:** Using large population cohort data from the UK Biobank, we employed logistic and Cox regression to explore the cross-sectional and longitudinal associations between dietary iron intake and mental and behavioral disorders due to use of tobacco. Additionally, we assessed the nonlinear relationship between dietary iron intake and mental and behavioral disorders due to use of tobacco using restricted cubic spline plots.

**Results:** The cross-sectional analysis included 50,991 participants. The logistic regression results indicated that dietary iron intake was negatively associated with mental and behavioral disorders due to use of tobacco. A total of 50,921 participants were included in the cohort study. The Cox regression results supported the protective effect of increased dietary iron intake against mental and behavioral disorders due to use of tobacco. The stratified and sensitivity analysis results were consistent with the main results. The restricted cubic spline plots showed a nonlinear relationship between dietary iron intake and mental and behavioral disorders due to use of tobacco. The risk reduction rate initially accelerated and then slowed in the total sample, the two age, and the male groups. In contrast, it declined rapidly at first and then leveled off in the female group.

**Conclusion:** This study found that dietary iron intake has a protective effect against mental and behavioral disorders due to use of tobacco, revealing a nonlinear association between the two. These findings offer valuable insights for the prevention and treatment of mental and behavioral disorders due to use of tobacco in the future.

**What is already known on this topic:** Existing research primarily focuses on tobacco as a risk factor for physical diseases. In contrast, the factors influencing mental and behavioral disorders due to use of tobacco have not been adequately explored. Furthermore, findings regarding the relationship between dietary iron intake and mental health in the general population are inconsistent, highlighting the need for this study to clarify the potential association between dietary iron intake and mental and behavioral disorders due to use of tobacco among smokers.

**What this study adds:** In this combined cross-sectional and longitudinal study, we assessed the association between dietary iron intake and mental and behavioral disorders due to use of tobacco using data from UK Biobank. We found that high dietary iron intake was protective against mental and behavioral disorders due to use of tobacco. In the fully adjusted model, the OR (95% CI) and HR (95% CI) for the highest intake group compared to the lowest intake group were 0.41 (0.18 - 0.98) and 0.50 (0.43 - 0.58), respectively. In addition, we found a similar L-shaped nonlinear association between dietary iron intake and mental and behavioral disorders due to use of tobacco utilizing restricted cubic spline plots.

**How this study might affect research, practice or policy:** Our study provides evidence of a negative association between dietary iron intake and mental and behavioral disorders due to use of tobacco. For groups that find it difficult to quit smoking, increasing iron intake to an appropriate level may alleviate the discomfort associated with mental and behavioral disorders due to use of tobacco. National mental health is crucial for every country, and this becomes especially important in a modern context where mental stress is increasingly recognized.

## 1 INTRODUCTION

The tobacco epidemic is one of the greatest public health threats that the world has ever faced[1]. The clear realization that tobacco is a risk factor for a variety of diseases, including lung cancer, attracts the vast majority of attention[2–6]. However, mental and behavioral disorders due to use of tobacco have rarely been studied. Mental and behavioral disorders due to the use of tobacco refer to dependence syndrome, harmful use, withdrawal state, and acute intoxication, etc.[7]. These disorders are characterized by strong urges to smoke, inability to control cessation, and withdrawal symptoms such as frustration, anxiety, and irritability[8]. There were more than 1 billion smokers worldwide in 2019, which is still increasing today[9]. According to the American Lung Association, 35% of smokers have behavioral health disorders[10]. Among those hospitalized for schizophrenia, depression, or bipolar disorder, approximately half of the deaths are due to smoking-related causes[11]. Therefore, it is critical to identify and manage the influence factors for mental and behavioral disorders due to use of tobacco.

Tobacco use causes nicotine to bind to and activate acetylcholine receptors in the body, which results in dopamine release[12]. However, repeated exposure to nicotine leads to desensitization of these receptors, resulting in decreased dopamine release and contributing to mental and behavioral disorders[13]. Iron, an essential nutrient, is closely related to physical and mental health[14–18]. One of the physiological roles of iron is to promote dopaminergic signaling by enhancing the expression of the D2 neurotransmitter, increasing dopamine release[19]. Based on this, we hypothesize that there is an association between dietary iron intake with mental and behavioral disorders due to use of tobacco.

Many studies have currently reported connections between iron and mental and behavioral disorders with inconsistent conclusions. For example, one study provided evidence of a negative correlation between circulating iron levels and depression and anxiety across different age and gender groups[20]. Similarly, another study found that dietary iron intake had a negative association with the risk of depression[21]. In contrast, one study indicated that excessive iron increased the risk of depression and anxiety[22]. Despite numerous investigations in the general population, there has been no research on the relationship between dietary iron intake and mental and behavioral disorders due to use of tobacco among smokers[23, 24].

Therefore, we conducted a cross-sectional and cohort study using data from the UK Biobank (UKB) to explore both the cross-sectional and longitudinal associations between dietary iron intake and mental and behavioral disorders due to use of tobacco. Additionally, we employed restricted cubic spline curves to clarify the dose-response relationship between dietary iron intake and mental and behavioral disorders due to use of tobacco.

## 2 METHODS

### 2.1 Study participant and design

The UKB is a large prospective population-based study that assessed approximately 500,000 participants at 22 assessment centers between 2006 and 2010, including detailed health data, genomic information, and lifestyle data for each individual. These assessment centers covered various settings in the UK, ensuring a broad distribution of all exposures to detect reliable generalized associations between baseline characteristics and health outcomes. Additionally, the UKB was combined with UK national datasets for longitudinal follow-up of individual information. The study has received ethical approval from the North West Multi-Centre Research Ethics Committee in the UK (reference number: 11/NW/0382, 16/NW10274, 21/NW/0157), and all participants provided written informed consent. The specific filtering process is illustrated in Figure 1.

**Figure 1.**
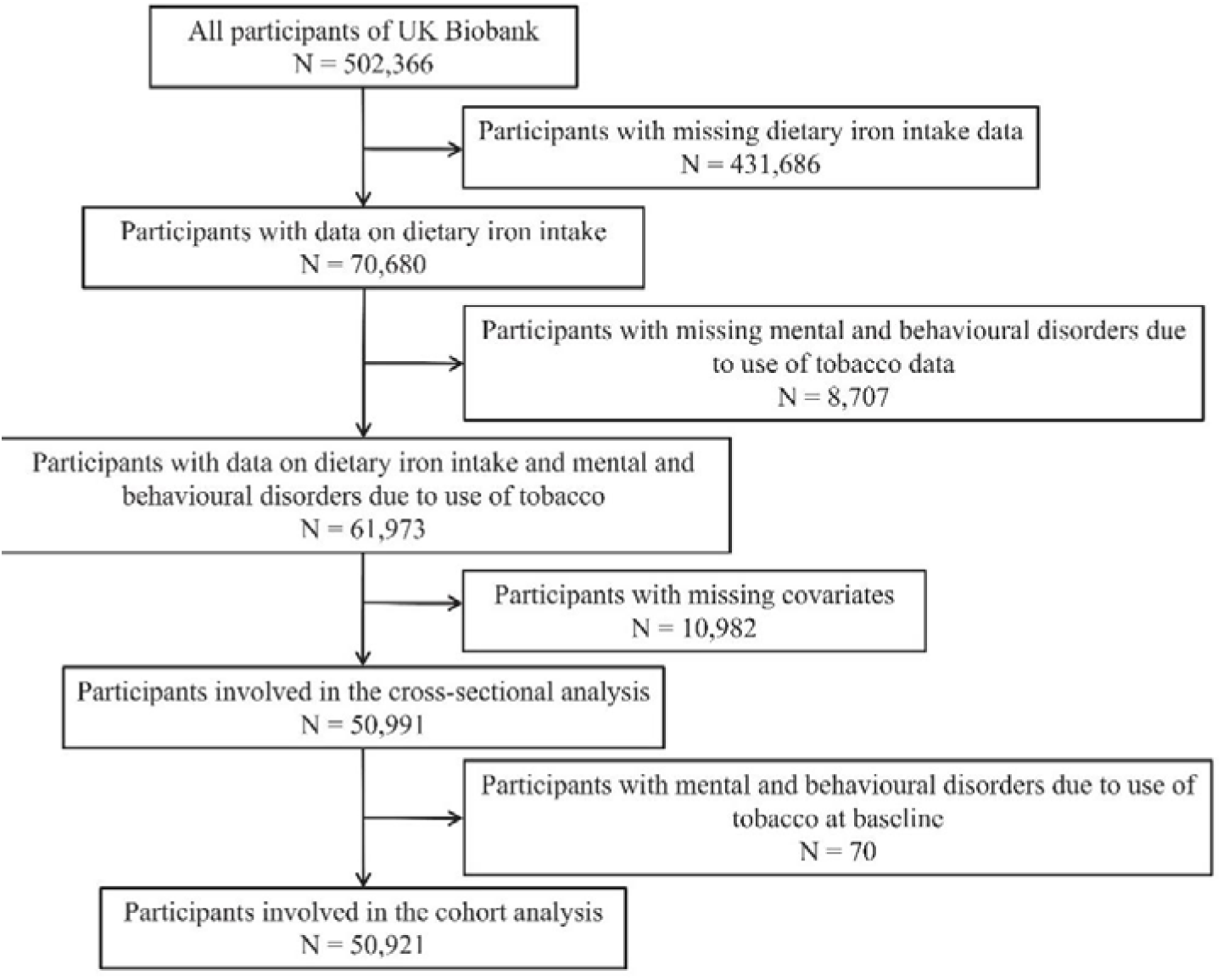
Flow diagram of study participant selection.

### 2.2 Exposure

Between April 2009 and September 2010, participants’ overall dietary intake information for the past 24 hours was collected at assessment centers in the UK using an online questionnaire called Oxford WebQ. This 24-hour dietary assessment tool is specifically designed for large prospective studies. It asks participants to provide dietary information using specified standard portion sizes within given food categories[25]. Statistical data indicated that Oxford WebQ correlated with most nutrients equal to or greater than interviewer-administered dietary information[26]. In the Oxford WebQ, the intake of food nutrients, including iron, was automatically calculated by multiplying the consumed portions by the predetermined quantity of each food and its corresponding nutrient content[27].

### 2.3 Outcome

The outcome of this study was mental and behavioral disorders due to use of tobacco, identified using the International Classification of Diseases, 10th edition (ICD-10) codes. The outcome code was F17 in ICD-10, including F17.0 (acute intoxication), F17.1 (harmful use), F17.2 (dependence syndrome), F17.3 (withdrawal state), F17.4 (withdrawal state with delirium), F17.7 (residual and late-onset psychotic disorder), F17.9 (unspecified mental and behavioral disorder), with more details available on the UKB website (https://www.ukbiobank.ac.uk/). New cases and the onset time were determined based on the first diagnosis recorded in the hospital inpatient database during the follow-up period. The date of death was obtained from the UK National Death Register. The follow-up period ended on the date of onset, date of death, date of loss to follow-up, or the date of observation termination (December 31, 2022), with the earliest occurrence taking precedence.

### 2.4 Covariates

Based on previous research, the following factors were included as covariates in the study: age, sex (male, female), ethnicity (White, other), body mass index (BMI), physical activity level (low, moderate, high), energy intake, Townsend Deprivation Index (TDI), education qualifications (“college or university degree and other professional qualifications” were categorized as “College and above,” while the remaining qualifications were categorized as “Below College”), alcohol consumption (never, previous, current), employment status (“employed in paid employment or self-employed” was categorized as “Full-time work”, while the remaining categories were classified as “Non-full-time work”), hypertension (yes, no), stroke (yes, no), and diabetes (yes, no)[28–36]. Physical activity levels were classified based on Metabolic Equivalent Task (MET) score data derived from the International Physical Activity Questionnaire (IPAQ) guidelines[37].

### 2.5 Statistical analysis

In descriptive studies, continuous variables were described by mean and standard deviation (SD) and categorical variables by numbers (N) and percentages (%). Dietary iron intake was divided into four quartiles (Q1: <25th percentile, Q2: 25th to 50th percentile, Q3: 50th to 75th percentile, and Q4: >75th percentile), with the lowest quartile as the reference group.

In cross-sectional studies, logistic regression models were employed to estimate the association between dietary iron intake and mental and behavioral disorders due to use of tobacco. Results were reported as odds ratios (OR) and 95% confidence intervals (CI).

In the cohort study, the Cox proportional hazards model was used to assess the longitudinal associations between dietary iron intake and mental and behavioral disorders due to use of tobacco. The time scale was the follow-up duration from baseline to the occurrence of outcomes, death, loss to follow-up, or the end of observation (whichever occurred first, measured in days). Results were reported as hazard ratios (HR) with 95%CI.

A total of four models were fitted in this study. Model 1 included no covariates. Model 2 adjusted for age, sex, ethnicity, education qualifications, employment status, and TDI. Model 3 further adjusted for BMI, physical activity level, energy intake, and alcohol consumption. Model 4 was additionally adjusted for hypertension, diabetes, and stroke.

To further evaluate the robustness of the cohort analysis results, we conducted the following sensitivity analyses: (1) Considering the chronic and long-term nature of the effects of dietary factors on mental and behavioral disorders in the cohort analysis, we excluded participants diagnosed with mental and behavioral disorders due to use of tobacco in the 2 years prior to follow-up. (2) Given the strong association between sleep and mental health, we adjusted for sleep duration[38]. A sleep duration of 7-8 hours was classified as normal sleep, while other durations were classified as abnormal sleep. (3) Participants with extreme iron intake were excluded (intake < first percentile and > ninety-ninth percentile). (4) Participants with a history of hypertension, diabetes, or stroke at baseline were excluded. Moreover, stratified analyses were performed according to sex, age, and BMI, while the interaction was examined using a Wald test. Age was divided into two groups: ≤60 years and >60 years, and BMI was divided into four groups: <18.5 kg/m^2^, ≥ 18.5 kg/m^2^ & <25 kg/m^2^, ≥25 kg/m^2^ & <30 kg/m^2^, ≥30 kg/m^2^.

Additionally, we treated dietary iron intake as a continuous variable and employed restricted cubic splines to examine the dose-response relationship between dietary iron intake and mental and behavioral disorders due to use of tobacco. Considering the varying iron requirements based on age and sex, we investigated the dose-response relationships separately for different gender and age groups.

Statistical analyses were performed in R version 4.2.3, and a *P*-value < 0.05 for a two-sided test was considered statistically significant.

## 3 RESULTS

### 3.1 Participant baseline characteristics

The cross-sectional analysis included 50,991 participants and a total of 50,921 participants were included in the cohort study. The baseline characteristics of study participants grouped by dietary iron intake are shown in Table 1. Compared to participants with the lowest dietary iron intake, those with the highest intake were more likely to be male, non-full-time workers, and individuals with diabetes. They tended to have lower BMI and TDI while exhibiting higher levels of physical activity, energy intake, education qualifications, and alcohol consumption.

**Table 1.**
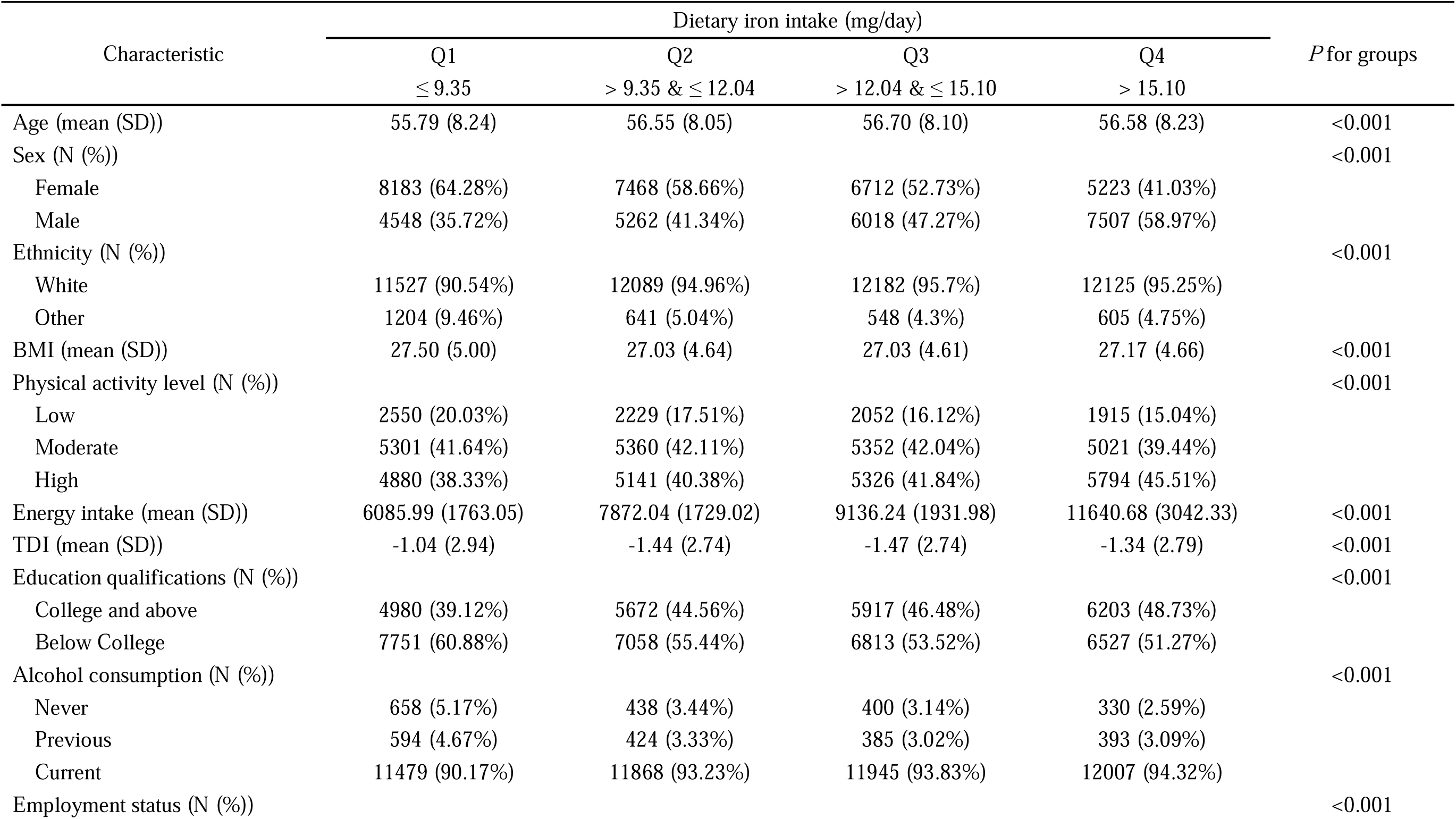
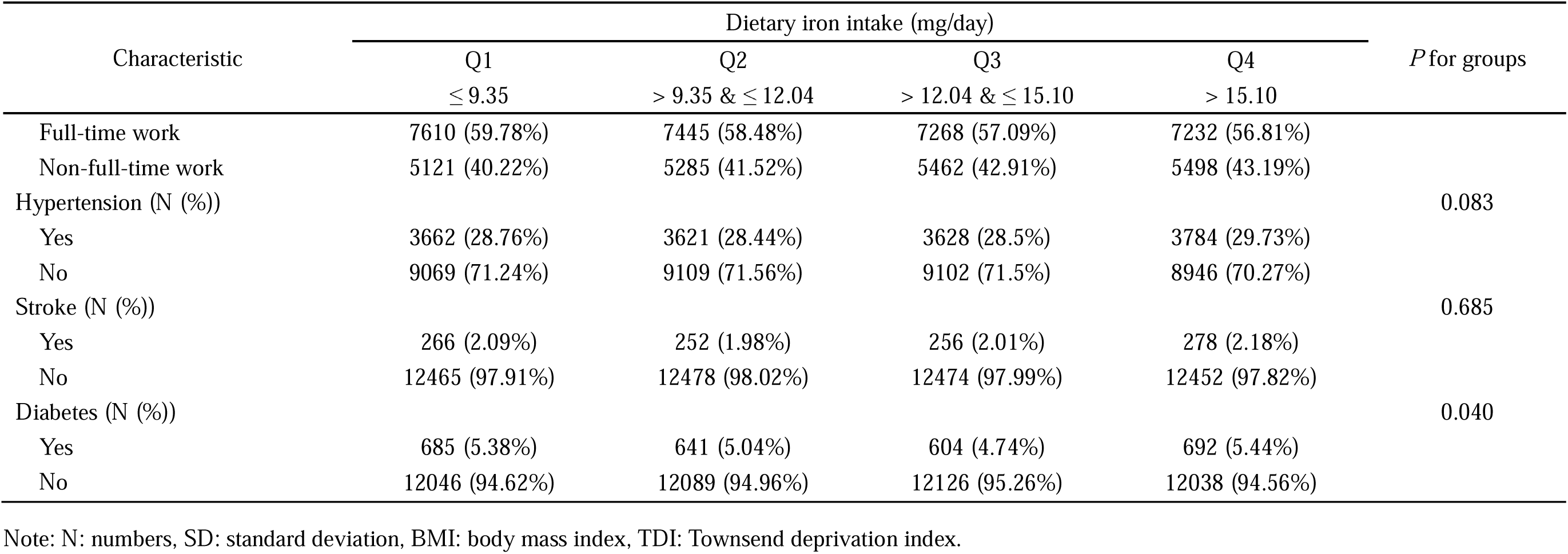
Baseline characteristics of the quartile of dietary iron intake.

### 3.2 Cross-sectional analysis

At baseline, there were 70 patients with mental and behavioral disorders due to use of tobacco. The results of the cross-sectional analysis are displayed in Table 2. According to the results, all four models showed that dietary iron intake was negatively associated with mental and behavioral disorders due to use of tobacco. The model 4 (adjusted for age, sex, ethnicity, BMI, physical activity level, energy intake, TDI, education qualifications, alcohol consumption, employment status, hypertension, stroke, and diabetes) indicated that compared to the Q1 group, OR (95%CI) for the Q2, Q3, and Q4 groups were 0.42 (0.21 - 0.83), 0.26 (0.12 - 0.60), and 0.41 (0.18 - 0.98), respectively.

**Table 2.** Association between dietary iron intake and mental and behavioral disorders due to use of tobacco in the cross-sectional study.

| Dietary iron intake | Model1 |  | Model2 |  | Model3 |  | Model4 |  |
| --- | --- | --- | --- | --- | --- | --- | --- | --- |
|  | OR (95%CI) | *P*-value | OR (95%CI) | *P*-value | OR (95%CI) | *P*-value | OR (95%CI) | *P*-value |
| Q1 | 1.00 (Ref.) | Ref. | 1.00 (Ref.) | Ref. | 1.00 (Ref.) | Ref. | 1.00 (Ref.) | Ref. |
| Q2 | 0.42 (0.22 - 0.80) | **0.008** | 0.44 (0.23 - 0.85) | **0.014** | 0.42 (0.21 - 0.83) | **0.013** | 0.42 (0.21 - 0.83) | **0.012** |
| Q3 | 0.29 (0.14 - 0.61) | **0.001** | 0.29 (0.14 - 0.62) | **0.001** | 0.27 (0.12 - 0.61) | **0.002** | 0.26 (0.12 - 0.60) | **0.001** |
| Q4 | 0.55 (0.30 - 0.99) | **0.046** | 0.51 (0.28 - 0.94) | **0.030** | 0.43 (0.18 - 1.01) | 0.053 | 0.41 (0.18 - 0.98) | **0.044** |

Note: Model 1 was not adjusted for covariates. Model 2 adjusted for age, sex, ethnicity, education qualifications, employment status, and TDI. Model 3 further adjusted for BMI, physical activity level, energy intake, and alcohol consumption. Model 4 further adjusted for hypertension, diabetes, and stroke.

### 3.3 Cohort analysis

During a median follow-up of 6.84 years, 1,404 new cases of mental and behavioral disorders due to use of tobacco were identified. The association between dietary iron intake and the risk of mental and behavioral disorders due to use of tobacco is presented in Table 3. The Cox proportional hazards model did not violate the proportional hazards assumption. According to the results, Model 4 indicated that the risk of mental and behavioral disorders due to use of tobacco was reduced in Q2 (HR: 0.60, 95%CI: 0.53 - 0.68, *P*: <0.001), Q3 (HR: 0.59, 95%CI: 0.52 - 0.67, *P*: <0.001), and Q4 (HR: 0.50, 95%CI: 0.43 - 0.58, *P*: <0.001) groups compared to the Q1 group, and this association was observed in Models 1, 2, and 3.

**Table 3.** Longitudinal association between dietary iron intake and mental and behavioral disorders due to use of tobacco.

| Dietary iron intake | Model1 |  | Model2 |  | Model3 |  | Model4 |  |
| --- | --- | --- | --- | --- | --- | --- | --- | --- |
|  | HR (95%CI) | *P*-value | HR (95%CI) | *P*-value | HR (95%CI) | *P*-value | HR (95%CI) | *P*-value |
| Q1 | 1.00 (Ref.) | Ref. | 1.00 (Ref.) | Ref. | 1.00 (Ref.) | Ref. | 1.00 (Ref.) | Ref. |
| Q2 | 0.60 (0.53 - 0.68) | **<0.001** | 0.67 (0.59 - 0.75) | **<0.001** | 0.60 (0.53 - 0.68) | **<0.001** | 0.60 (0.53 - 0.68) | **<0.001** |
| Q3 | 0.59 (0.52 - 0.67) | **<0.001** | 0.70 (0.63 - 0.79) | **<0.001** | 0.59 (0.52 - 0.67) | **<0.001** | 0.59 (0.52 - 0.67) | **<0.001** |
| Q4 | 0.50 (0.43 - 0.59) | **<0.001** | 0.71 (0.63 - 0.79) | **<0.001** | 0.50 (0.43 - 0.59) | **<0.001** | 0.50 (0.43 - 0.58) | **<0.001** |

Note: Model 1 was not adjusted for covariates. Model 2 adjusted for age, sex, ethnicity, education qualifications, employment status, and TDI. Model 3 further adjusted for BMI, physical activity level, energy intake, and alcohol consumption. Model 4 further adjusted for hypertension, diabetes, and stroke.

### 3.4 Stratified analysis

The stratified analysis results are shown in Supplementary Table 1. When stratified by age, both age groups resulted in a reduced risk of mental and behavioral disorders due to use of tobacco as dietary iron intake increased. In the ≤60 age group, the HR (95% CI) was 0.58 (0.50 - 0.67), 0.57 (0.49 - 0.67), and 0.45 (0.37 - 0.55) for the Q2, Q3, and Q4 groups, respectively, compared to the Q1 group. In the>60 age group, HR (95% CI) was 0.62 (0.51 - 0.76), 0.60 (0.49 - 0.75), and 0.56 (0.44 - 0.73) in Q2, Q3, and Q4 groups respectively compared to the Q1 group.

When analyzed by sex, the male and female results were consistent with the main analysis findings. In females, the HR (95% CI) for the Q2, Q3, and Q4 groups compared to the Q1 group were 0.63 (0.53 - 0.75), 0.60 (0.50 - 0.73), and 0.59 (0.47 - 0.75), respectively. In males, the HR (95% CI) for the Q2, Q3, and Q4 groups compared to the Q1 group were 0.58 (0.49 - 0.68), 0.58 (0.49 - 0.69), and 0.45 (0.37 - 0.55), respectively.

When stratifying by BMI, only the group with BMI < 18.5 kg/m^2^ did not yield statistically significant results. This might have been due to the small sample size of this subgroup (N = 263). In the group with BMI ≥ 18.5 kg/m² & < 25 kg/m², the HR (95% CI) for the Q2, Q3, and Q4 groups compared to the Q1 group were 0.50 (0.40 - 0.62), 0.47 (0.38 - 0.59), and 0.37 (0.29 - 0.49), respectively. In the group with BMI ≥ 25 kg/m² & < 30 kg/m², the HR (95% CI) for the Q2, Q3, and Q4 groups were 0.62 (0.51 - 0.74), 0.61 (0.50 - 0.74), and 0.50 (0.40 - 0.64), respectively. In the group with BMI ≥ 30 kg/m², the HR (95% CI) for the Q2, Q3, and Q4 groups were 0.72 (0.57 - 0.90), 0.76 (0.59 - 0.97), and 0.68 (0.51 - 0.92), respectively.

Furthermore, no statistically significant differences were observed between the subgroups.

### 3.4 Sensitivity analysis

After excluding individuals diagnosed within the first two years of follow-up, the association between higher dietary iron intake and a lower risk of mental and behavioral disorders due to use of tobacco remained unchanged (Supplementary Table 2). In Model 4, In Model 4, groups Q2 (HR: 0.60, 95%CI: 0.53-0.68, *P* < 0.001), Q3 (HR: 0.59, 95%CI: 0.52-0.67, *P* < 0.001) and Q4 (HR: 0.52, 95%CI: 0.44-0.61, *P* < 0.001) had reduced risk compared to group Q1.

Adjusting for sleep duration did not significantly change the main analysis results (Supplementary Table 3). In Model 4, compared to group Q1, the risk was reduced in Q2 (HR: 0.61, 95%CI: 0.54 - 0.68, *P* < 0.001), Q3 (HR: 0.60, 95%CI: 0.53 - 0.67, *P* < 0.001), and Q4 (HR: 0.51, 95%CI: 0.43 - 0.59, *P* < 0.001) groups.

Additionally, after excluding participants with extreme dietary iron intake, the findings remained consistent with the main analysis (Supplementary Table 4). In Model 4, groups Q2 (HR: 0.62, 95%CI: 0.55-0.70, *P* < 0.001), Q3 (HR: 0.60, 95%CI: 0.52-0.68, *P* < 0.001) and Q4 (HR: 0.49, 95%CI: 0.42-0.57, *P* < 0.001) had reduced risk compared to group Q1.

Finally, excluding participants with hypertension, stroke, and diabetes did not alter the association (Supplementary Table 5). In Model 3, compared to group Q1, the risk was reduced in Q2 (HR: 0.59, 95%CI: 0.51 - 0.68, *P* < 0.001), Q3 (HR: 0.56, 95%CI: 0.48 - 0.65, *P* < 0.001), and Q4 (HR: 0.47, 95%CI: 0.39 - 0.56, *P* < 0.001) groups.

These sensitivity analysis results confirmed the robustness of the study findings.

### 3.6 Restricted cubic spline

The results of the restricted cubic spline analysis indicated a similar L-shaped nonlinear relationship between dietary iron intake and mental and behavioral disorders due to use of tobacco. As dietary iron intake increased, the risk of these disorders decreased. A significant risk reduction was observed when intake was approximately between 0 to 12 mg/day. However, the risk reduction rate slowed when the intake exceeded about 12 mg/day (Figure 2).

**Figure 2.**
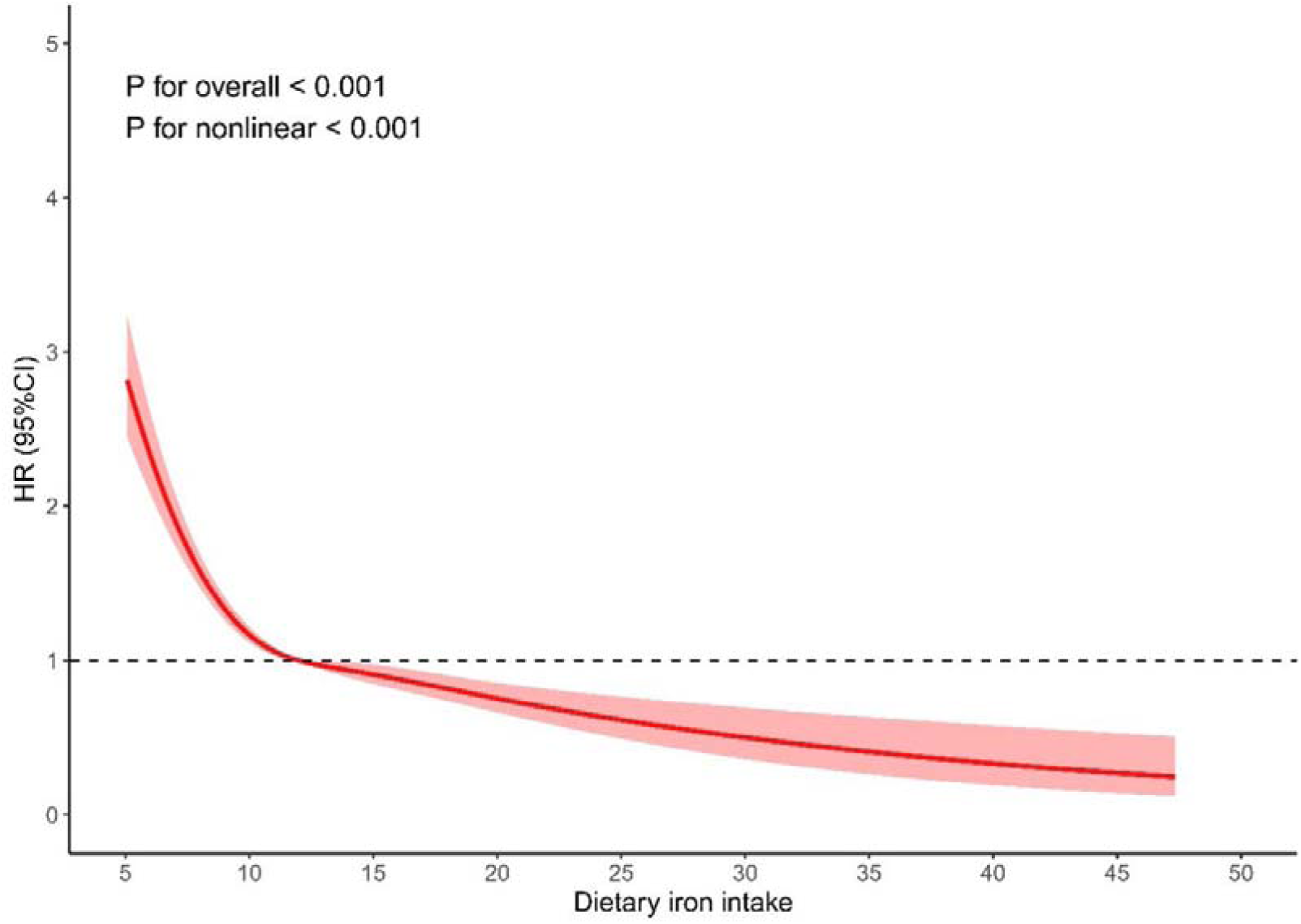
Restricted cubic spline plots of dietary iron intake and mental and behavioral disorders due to use of tobacco.

When stratified by age, the trend in risk changes was similar to what was previously described. The inflection point for the ≤60 years and the >60 years groups was around 12 mg/day (Supplementary Figures 1a and 1b).

When stratified by sex, the trend in risk change for males followed a similar pattern to that previously described, with an inflection point at approximately 13 mg/day (Supplementary Figure 1c). In females, however, an L-shape was observed between dietary iron intake and mental and behavioral disorders due to use of tobacco. A significant risk reduction occurred when intake was around 0 to 11 mg/day. When intake exceeded approximately 11 mg/day, the risk reduction rate began to plateau (Supplementary Figure 1d).

## 4 DISCUSSION

In this study, we conducted logistic regression and Cox regression analyses based on the UKB population sample, finding that higher dietary iron intake was associated with a lower risk of mental and behavioral disorders due to use of tobacco. Stratified analysis results supported the protective effect of dietary iron intake on mental and behavioral disorders due to use of tobacco. The sensitivity analysis results were consistent with the main analysis results. Additionally, the restricted cubic spline analysis revealed a nonlinear relationship between dietary iron intake and mental and behavioral disorders due to use of tobacco. In the overall population, both age groups and the male group, the risk of increasing dietary iron intake initially decreased rapidly before slowing down. For females, the trend in risk change was characterized by a rapid initial decline followed by stabilization.

In this study, we observed that higher dietary iron intake was associated with a reduced risk of mental and behavioral disorders due to use of tobacco. Although there is no epidemiological evidence linking dietary iron intake to mental and behavioral disorders due to use of tobacco, a large number of previous studies have explored the relationship between iron and mental and behavioral disorders. It is evident that mental and behavioral disorders due to use of tobacco are a subset of this broader category. For example, one clinical study found that iron deficiency significantly increased the risk of both psychiatric comorbidity and individual psychiatric disorders[39]. Results from another study supported the association of iron deficiency with depressive symptoms[40]. A clinical study revealed that low iron levels were associated with a significantly higher risk of developing mental disorders[41]. In addition, animal experiments found that rats fed an iron-deficient diet exhibited more anxiety-like behaviors than control rats, supporting the findings of observational studies in humans[42]. These findings supported our results to some extent.

Mental and behavioral disorders due to use of tobacco are primarily driven by nicotine dependence[43]. Nicotine is a sympathomimetic stimulant that binds to nicotinic acetylcholine receptors in the brain[44, 45]. It releases dopamine and other neurotransmitters, such as norepinephrine, which induce feelings of pleasure[46]. Repeated exposure to nicotine increases the number of nicotinic acetylcholine receptors, altering the brain’s reward system and raising the threshold for pleasure[46, 47]. When these receptors are not bound to nicotine, individuals may experience mental and behavioral disorders such as inattention, restlessness, and irritability[43]. Iron plays a crucial role in the synthesis and release of dopamine. On the one hand, iron can increase dopamine release by promoting the work of dopamine transporter proteins[48]. On the other hand, iron can enhance dopaminergic signaling by increasing the affinity and expression of D2 neurotransmitters[49]. In addition, iron is an essential cofactor for tyrosine hydroxylase, the rate-limiting enzyme in dopamine synthesis[50]. Sufficient iron levels can enhance tyrosine hydroxylase activity, thereby increasing dopamine production and alleviating mental disorders[51].

This study had several strengths. Firstly, it represented the first exploration of the association between dietary iron intake and mental and behavioral disorders due to use of tobacco. In addition, we explored the L-shaped and similar L-shape association between dietary iron intake and mental and behavioral disorders due to use of tobacco using restricted cubic spline plots. Secondly, we combined both cross-sectional and longitudinal studies, verifying the results from the cross-sectional analysis with cohort studies to ensure reliability. Thirdly, we utilized a large population cohort from the UKB, enhancing the representativeness of our sample. Finally, we comprehensively included various covariates and conducted multiple sensitivity and stratified analyses to assess the robustness of our findings.

We acknowledged several limitations. Firstly, dietary iron intake information was obtained from participants’ online questionnaires, which might introduce recall bias. Secondly, the prolonged data collection period of the UKB prevented us from distinguishing the effects of short-term versus long-term dietary iron intake on mental and behavioral disorders due to use of tobacco. Thirdly, the study was ultimately observational, preventing us from drawing causal conclusions. Lastly, although we had attempted to adjust for numerous confounding factors, the influence of unknown factors may persist.

## 5. CONCLUSION

In conclusion, we found a stable, protective effect of dietary iron intake against mental and behavioral disorders due to use of tobacco and a nonlinear relationship between dietary iron intake and mental and behavioral disorders due to use of tobacco. Our findings may provide valuable insights into how individuals can reduce mental and behavioral disorders due to use of tobacco by appropriately increasing dietary iron intake. Meanwhile, it offers reliable clues for future public health policy development.

## Acknowledgments

We thank all the staff at the UKB. This research has been conducted using the UKB Resource under Application Number 95715

## Contributors

XQ contributed to the conception, design, acquisition and interpretation of data, performed all statistical analyses and drafted and critically revised the manuscript. RZ, QZ, and JL contributed to the design, analysis and interpretation of results. TW and WW contributed to analysis and interpretation of results and critically revised the manuscript. HZ makes critical revisions to important intellectual content. DZ contributed to the conception, design and acquisition of data and critically revised the manuscript. All authors gave their final approval and agreed to be accountable for all aspects of the work.

## Patient consent for publication

Not required.

## Ethics approval

Ethical review for this study was obtained from the North West Multi-Centre Research Ethics Committee in the UK (reference number: 11/NW/0382, 16/NW10274, 21/NW/0157).

## Data availability statement

The data supporting the findings of this study are publicly available at UK Biobank (https://www.ukbiobank.ac.uk).

