## Supplementary Tables 1-5 and Supplementary Figure 1 for "Dietary Iron Intake and Mental and Behavioral Disorders Due to Use of Tobacco: A Cross-Sectional and Longitudinal Study Based on the UK Biobank": Supplementary Material.docx

**Supplementary material Contents**

**Supplementary Table 1** Stratified analysis of associations between dietary iron intake and mental and behavioral disorders due to use of tobacco based on baseline age, sex, and BMI (Model 4).

| Characters | Q2 | | |  | Q3 | | |  | Q4 | | |
| --- | --- | --- | --- | --- | --- | --- | --- | --- | --- | --- | --- |
|  | HR (95%CI) | *P*-value | *P*-value for interaction |  | HR (95%CI) | *P*-value | *P*-value for interaction |  | HR (95%CI) | *P*-value | *P*-value for interaction |
| Age |  |  |  |  |  |  |  |  |  |  |  |
| ≤ 60 years | 0.58 (0.50 - 0.67) | **<0.001** | 0.612 |  | 0.57 (0.49 - 0.67) | **<0.001** | 0.355 |  | 0.45 (0.37 - 0.55) | **<0.001** | 0.060 |
| > 60 years | 0.62 (0.51 - 0.76) | **<0.001** |  |  | 0.60 (0.49 - 0.75) | **<0.001** |  |  | 0.56 (0.44 - 0.73) | **<0.001** |  |
| Sex |  |  |  |  |  |  |  |  |  |  |  |
| Female | 0.63 (0.53 - 0.75) | **<0.001** | 0.638 |  | 0.60 (0.50 - 0.73) | **<0.001** | 0.740 |  | 0.59 (0.47 - 0.75) | **<0.001** | 0.247 |
| Male | 0.58 (0.49 - 0.68) | **<0.001** |  |  | 0.58 (0.49 - 0.69) | **<0.001** |  |  | 0.45 (0.37 - 0.55) | **<0.001** |  |
| BMI |  |  |  |  |  |  |  |  |  |  |  |
| <18.5 kg/m^2^ | 0.77 (0.22 - 2.63) | 0.671 | 0.096 |  | 0.20 (0.02 - 1.95) | 0.167 | 0.228 |  | 0.87 (0.16 - 4.57) | 0.866 | 0.351 |
| ≥18.5 kg/m^2^ & <25 kg/m^2^ | 0.50 (0.40 - 0.62) | **<0.001** |  |  | 0.47 (0.38 - 0.59) | **<0.001** |  |  | 0.37 (0.29 - 0.49) | **<0.001** |  |
| ≥25 kg/m^2^ & <30 kg/m^2^ | 0.62 (0.51 - 0.74) | **<0.001** |  |  | 0.61 (0.50 - 0.74) | **<0.001** |  |  | 0.50 (0.40 - 0.64) | **<0.001** |  |
| ≥30 kg/m^2^ | 0.72 (0.57 - 0.90) | **0.005** |  |  | 0.76 (0.59 - 0.97) | **0.028** |  |  | 0.68 (0.51 - 0.92) | **0.013** |  |

Note: BMI: Body mass index. Model 4 adjusted for age, sex, ethnicity, education qualifications, employment status, TDI, BMI, physical activity level, energy intake, alcohol consumption, hypertension, diabetes, and stroke.

**Supplementary Table 2** Longitudinal associations between dietary iron intake and mental and behavioral disorders due to use of tobacco after excluding participants with the onset of disease in the first two years of follow-up.

| Model | Q2 | |  | Q3 | |  | Q4 | |
| --- | --- | --- | --- | --- | --- | --- | --- | --- |
|  | HR (95%CI) | *P*-value |  | HR (95%CI) | *P*-value |  | HR (95%CI) | *P*-value |
| Model1 | 0.63 (0.56 - 0.72) | **<0.001** |  | 0.68 (0.61 - 0.77) | **<0.001** |  | 0.75 (0.67 - 0.85) | **<0.001** |
| Model2 | 0.66 (0.59 - 0.75) | **<0.001** |  | 0.70 (0.62 - 0.79) | **<0.001** |  | 0.73 (0.65 - 0.82) | **<0.001** |
| Model3 | 0.60 (0.53 - 0.68) | **<0.001** |  | 0.59 (0.52 - 0.67) | **<0.001** |  | 0.52 (0.44 - 0.61) | **<0.001** |
| Model4 | 0.60 (0.53 - 0.68) | **<0.001** |  | 0.59 (0.52 - 0.67) | **<0.001** |  | 0.52 (0.44 - 0.61) | **<0.001** |

Note: Model 1 was not adjusted for covariates. Model 2 adjusted for age, sex, ethnicity, education qualifications, employment status, and TDI. Model 3 further adjusted for BMI, physical activity level, energy intake, and alcohol consumption. Model 4 further adjusted for hypertension, diabetes, and stroke.

**Supplementary Table 3** Longitudinal associations between dietary iron intake and mental and behavioral disorders due to use of tobacco after correcting for sleep duration effects.

| Model | Q2 | |  | Q3 | |  | Q4 | |
| --- | --- | --- | --- | --- | --- | --- | --- | --- |
|  | HR (95%CI) | P-value |  | HR (95%CI) | P-value |  | HR (95%CI) | P-value |
| Model1 | 0.64 (0.57 - 0.72) | **<0.001** |  | 0.69 (0.62 - 0.78) | **<0.001** |  | 0.74 (0.66 - 0.82) | **<0.001** |
| Model2 | 0.68 (0.60 - 0.76) | **<0.001** |  | 0.71 (0.64 - 0.80) | **<0.001** |  | 0.71 (0.64 - 0.80) | **<0.001** |
| Model3 | 0.61 (0.54 - 0.69) | **<0.001** |  | 0.60 (0.53 - 0.68) | **<0.001** |  | 0.51 (0.44 - 0.59) | **<0.001** |
| Model4 | 0.61 (0.54 - 0.68) | **<0.001** |  | 0.60 (0.53 - 0.67) | **<0.001** |  | 0.51 (0.43 - 0.59) | **<0.001** |

Note: Model 1 adjusted for sleep duration. Model 2 was further adjusted for age, sex, ethnicity, education qualifications, employment status, TDI, and sleep duration. Model 3 was further adjusted for BMI, physical activity level, energy intake, alcohol consumption, and sleep duration. Model 4 was further adjusted for hypertension, diabetes, stroke, and sleep duration.

**Supplementary Table 4** Longitudinal associations between dietary iron intake and mental and behavioral disorders due to use of tobacco after excluding participants with extreme dietary iron intake.

| Model | Q2 | |  | Q3 | |  | Q4 | |
| --- | --- | --- | --- | --- | --- | --- | --- | --- |
|  | HR (95%CI) | *P*-value |  | HR (95%CI) | *P*-value |  | HR (95%CI) | *P*-value |
| Model1 | 0.67 (0.60 - 0.75) | **<0.001** |  | 0.72 (0.64 - 0.81) | **<0.001** |  | 0.75 (0.67 - 0.84) | **<0.001** |
| Model2 | 0.70 (0.62 - 0.79) | **<0.001** |  | 0.74 (0.66 - 0.83) | **<0.001** |  | 0.72 (0.64 - 0.81) | **<0.001** |
| Model3 | 0.62 (0.55 - 0.70) | **<0.001** |  | 0.60 (0.53 - 0.68) | **<0.001** |  | 0.49 (0.42 - 0.58) | **<0.001** |
| Model4 | 0.62 (0.55 - 0.70) | **<0.001** |  | 0.60 (0.52 - 0.68) | **<0.001** |  | 0.49 (0.42 - 0.57) | **<0.001** |

Note: Model 1 was not adjusted for covariates. Model 2 adjusted for age, sex, ethnicity, education qualifications, employment status, and TDI. Model 3 further adjusted for BMI, physical activity level, energy intake, and alcohol consumption. Model 4 further adjusted for hypertension, diabetes, and stroke.

**Supplementary Table 5** Longitudinal associations between dietary iron intake and mental and behavioral disorders due to use of tobacco after excluding participants with hypertension, stroke, and diabetes.

| Model | Q2 | |  | Q3 | |  | Q4 | |
| --- | --- | --- | --- | --- | --- | --- | --- | --- |
|  | HR (95%CI) | *P*-value |  | HR (95%CI) | *P*-value |  | HR (95%CI) | *P*-value |
| Model1 | 0.64 (0.55 - 0.73) | **<0.001** |  | 0.67 (0.58 - 0.77) | **<0.001** |  | 0.74 (0.65 - 0.85) | **<0.001** |
| Model2 | 0.67 (0.58 - 0.77) | **<0.001** |  | 0.69 (0.60 - 0.80) | **<0.001** |  | 0.71 (0.62 - 0.82) | **<0.001** |
| Model3 | 0.59 (0.51 - 0.68) | **<0.001** |  | 0.56 (0.48 - 0.65) | **<0.001** |  | 0.47 (0.39 - 0.56) | **<0.001** |

Note: Model 1 was not adjusted for covariates. Model 2 adjusted for age, sex, ethnicity, education qualifications, employment status, and TDI. Model 3 further adjusted for BMI, physical activity level, energy intake, and alcohol consumption.

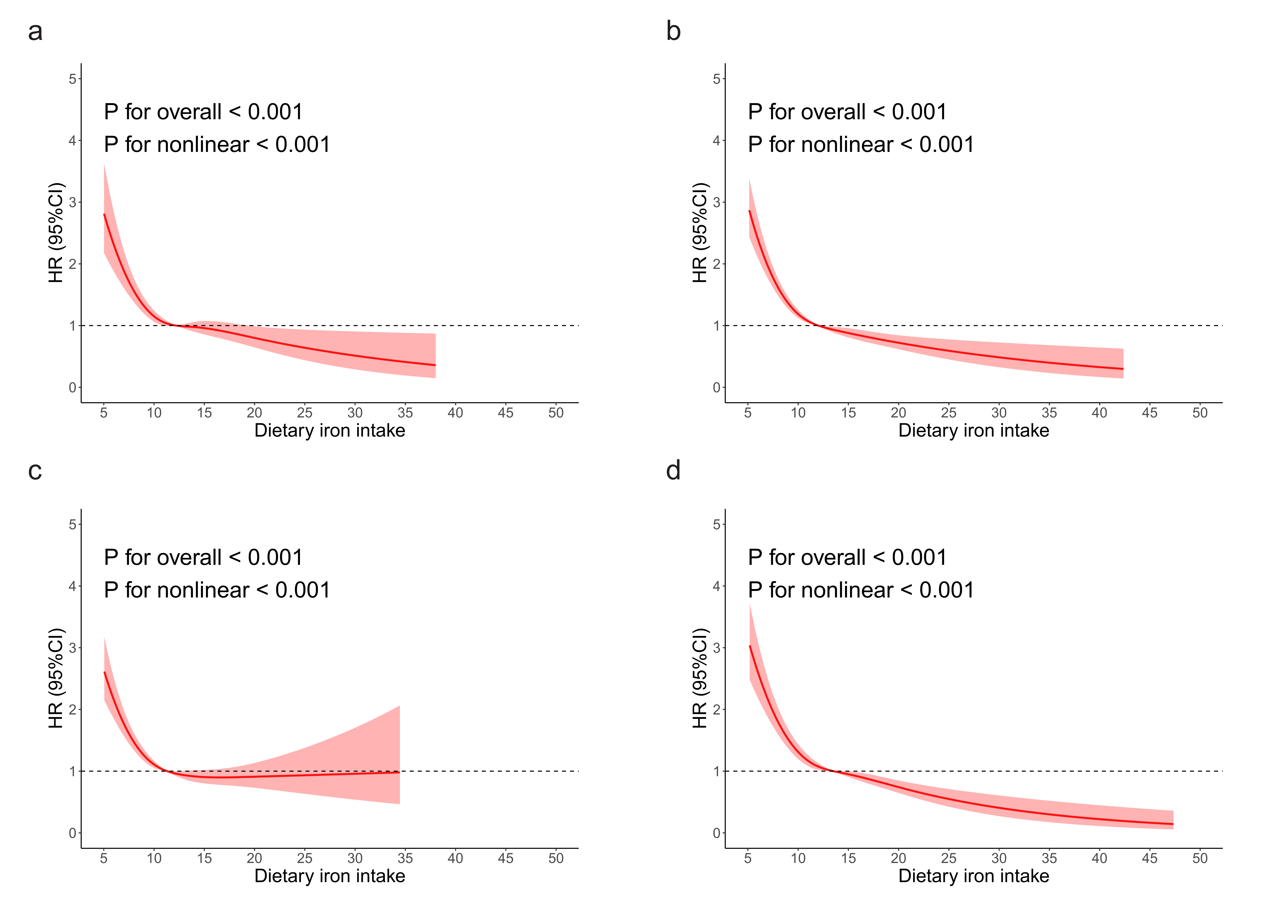

**Supplementary Figure 1** Restricted cubic spline plots of dietary iron intake and mental and behavioral disorders due to use of tobacco stratified by sex or age.

Note: a: ≤ 60 years, b: > 60 years, c: female, d: male.
